# A randomized, double-blind, Phase 1 study of IN-006, an inhaled antibody treatment for COVID-19

**DOI:** 10.1101/2022.08.17.22278748

**Authors:** Thomas R. Moench, Lakshmi Botta, Brian Farrer, Jason D. Lickliter, Hyunah Kang, Yoona Park, Cheolmin Kim, Marshall Hoke, Miles Brennan, Morgan D. McSweeney, Zachary Richardson, John B. Whelan, Jong Moon Cho, Soo Young Lee, Frances Faurot, Jeff Hutchins, Samuel K. Lai

## Abstract

**Rationale:** Although COVID-19 is predominantly a respiratory tract infection, current antibody treatments are administered by systemic dosing. We hypothesize that inhaled delivery of a muco-trapping monoclonal antibody would provide a more effective and convenient treatment for COVID-19.

**Objective:** We investigated the safety, tolerability, and pharmacokinetics of IN-006, a reformulation of regdanvimab, an approved intravenous treatment for COVID-19, for nebulized delivery by a handheld nebulizer.

**Methods:** A Phase 1 study was conducted in healthy volunteers. Study staff and participants were blinded to treatment assignment, except for pharmacy staff preparing the study drug. The primary outcomes were safety and tolerability. Exploratory outcomes were pharmacokinetic measurements of IN-006 in nasal fluid and serum.

**Results:** Twenty-three participants were enrolled and randomized across two single dose and one multiple dose cohorts. There were no serious adverse events (SAEs). All enrolled participants completed the study without treatment interruption or discontinuation. All treatment-emergent adverse events were transient, non-dose dependent, and were graded mild to moderate in severity. Nebulization was well tolerated and completed in a mean of 6 minutes in the high dose group. Mean nasal fluid concentrations of IN-006 in the multiple dose cohort were 921 µg/g of nasal fluid at 30 minutes after dosing and 5.8 µg/g at 22 hours. Mean serum levels in the multiple dose cohort peaked at 0.55 µg/mL at 3 days after the final dose.

**Conclusions:** IN-006 was well-tolerated and achieved concentrations in the respiratory tract orders of magnitude above its inhibitory concentration. These data support further clinical development of IN-006.

**Registration:** Australian New Zealand Clinical Trials Registry: ACTRN12621001235897

## Introduction

SARS-CoV-2 [1-3], like many viruses that cause acute respiratory infections (ARIs), infects cells almost exclusively via the apical (luminal) side of the airway epithelium and also primarily buds from infected cells via the apical surface [4-6]. Progeny virus must then travel through airway mucus to reach uninfected epithelial cells as the infection spreads from the upper respiratory tract (URT) to the lower respiratory tract (LRT) and the deep lung [7-9]. Neutralizing monoclonal antibodies (mAbs) must therefore reach a high enough concentration in the airway lumen to effectively neutralize the virus and halt the infection.

mAbs distribute very poorly and slowly from the blood into the respiratory tract fluids, with concentrations in the airways that are orders of magnitude lower than those in the serum following intravenous (IV) or intramuscular (IM) administration [10-12]. Despite these limitations, the clinical experience to date has shown that IV-administered mAbs against SARS-CoV-2 can be effective in treating infected individuals at high risk of severe COVID-19 when given early in the course of the infection [8, 9, 13], implying that sufficient amounts of mAb can distribute into the lung lumen. Nevertheless, high doses of mAb are generally required if given IV, reducing the number of treatment courses available from a given supply of drug. Delayed distribution into the lung also limits the treatment window for preventing severe COVID-19 [12].

Nebulization has been used to conveniently deliver protein therapeutics (e.g., Pulmozyme) directly to the lungs. Importantly, direct inhaled delivery can achieve far higher concentrations of drugs in the lungs than can be achieved by IV or IM administration, within minutes [14]. Since the pattern of deposition of nebulized drugs along the respiratory tract is largely determined by the aerosol particle size [15], it is possible to use a nebulizer that generates a broad aerosol size distribution to deliver drug throughout the entire respiratory tract, from the nasal turbinates in the URT, to conducting airways in the LRT, to the deep lung. Thus, nebulized delivery is likely the fastest method to achieve high, inhibitory concentrations of mAb in the airway fluids along the entire respiratory tract. Nebulization also enables convenient self-dosing at home, reducing the burden on patients and on the healthcare infrastructure compared to systemic delivery.

We are developing IN-006, a reformulation of regdanvimab for nebulized delivery, as an inhaled treatment for COVID-19. Regdanvimab, an IV-dosed human IgG_1_ mAb directed against the SARS-COV-2 spike protein receptor binding domain (RBD), is approved in the European Union for adults with COVID-19 who do not require supplemental oxygen and who are at increased risk of progression to severe COVID-19. We previously reported the results from preclinical activities that supported a first-in-human study of inhaled IN-006 [14], including muco-trapping of SARS-CoV-2 virions, GLP nebulization stability studies, and excellent tolerability in GLP inhaled toxicology and GLP tissue cross reactivity experiments. The current report describes a Phase 1 study designed to assess the safety, tolerability, and nasal fluid and serum pharmacokinetics of nebulized IN-006 in healthy adults.

## Methods

### Clinical Study Design and Participants

This double-blind, placebo-controlled, first-in-human, ascending-dose pharmacokinetic and safety study was conducted in a Phase 1 unit in Melbourne, Australia. The study was carried out according to the International Council for Harmonisation Good Clinical Practice guidelines and in compliance with local regulatory requirements and was approved by The Alfred Hospital Office of Ethics and Research Governance, Melbourne, VIC, Australia. This study was prospectively registered in the Australian New Zealand Clinical Trials Registry (ACTRN12621001235897).

Informed consent was obtained in advance of all study-related procedures. Eligible participants were enrolled sequentially into three cohorts: a single low dose cohort (30 mg), a single high-dose cohort (90 mg), and a multiple high-dose cohort (seven daily 90 mg doses). For each single dose cohort, a sentinel pair (with one active and one placebo recipient) was initially dosed, followed by a two-day safety monitoring period prior to the dosing of the remainder of the cohort. Advancing to subsequent cohorts was done after review of safety parameters seven days after final dosing of the preceding cohort. Figure 1 provides a diagram of the process flow, study structure, and times of pharmacokinetic evaluations.

**Figure 1.**
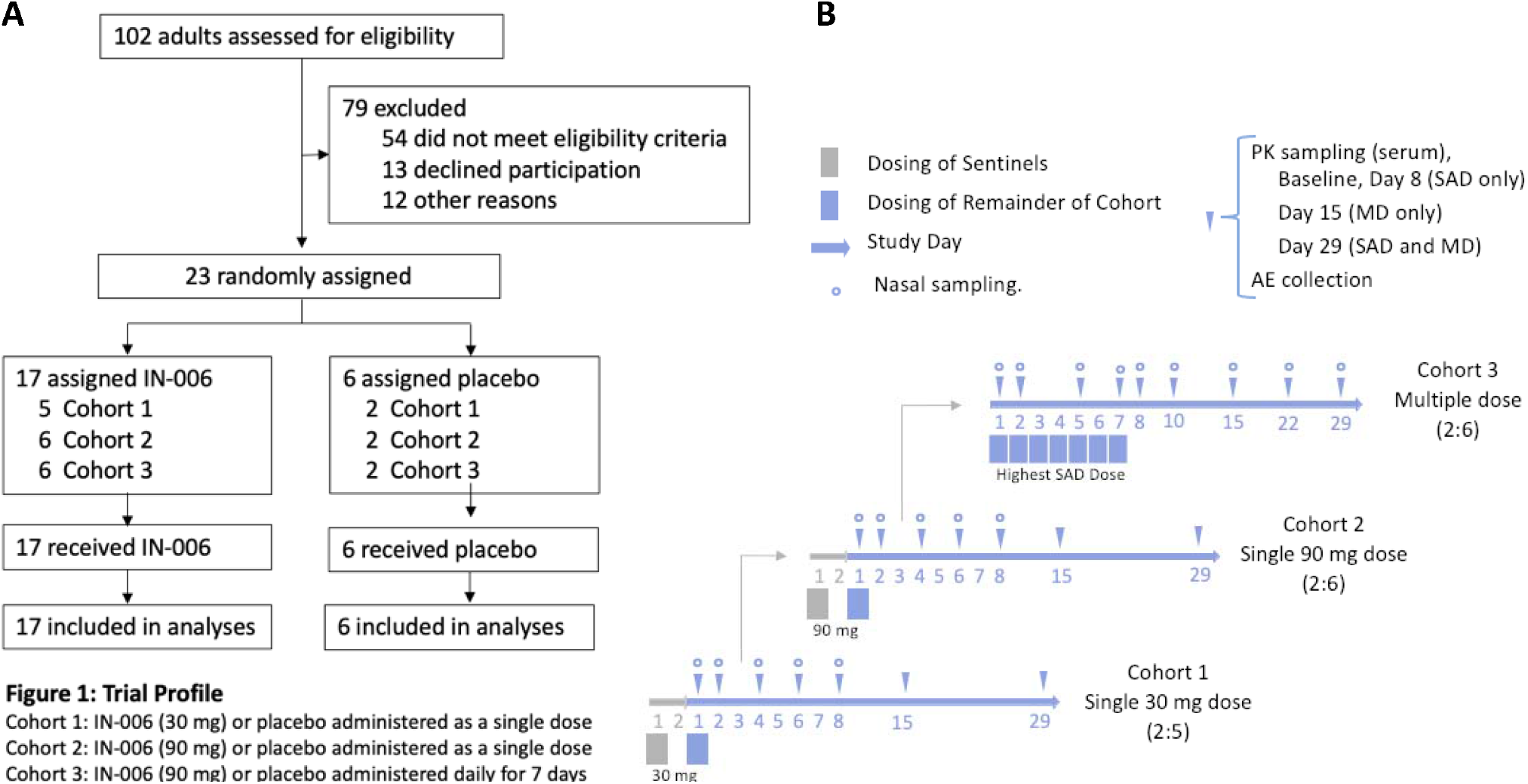
INH-006-01 clinical trial **(A)** process flow, and **(B)** study structure and times of sample collection for pharmacokinetic evaluations.

### Inclusion and Exclusion Criteria

Eligibility criteria required that participants be adults 18-55 years of age with a body-mass index of 18-32 kg/m^2^ who were in good health as judged by medical history, physical exam, clinical chemistry and hematology assessments, electrocardiogram, forced expiratory volume in one second (FEV_1_) ≥ 90% predicted, and negative serology for HBsAg, HCV, and HIV antibodies. Participants were required to be non- or light smokers. The FEV_1_ threshold was changed to ≥ 80% predicted after enrolling the first 7 participants.

Participants were excluded for known or suspected symptomatic viral infection or signs of active pulmonary infection or pulmonary inflammatory conditions within 14 days of dosing initiation, a history of airway hyperresponsiveness, angioedema, anaphylaxis, or a positive alcohol breathalyzer test and/or urine drug screen for substances of abuse.

During recruitment of the 7 participants comprising the first single dose cohort, participants who had received a COVID-19 vaccine were excluded. However, due to rapidly increasing local vaccine availability and uptake, this criterion was modified to exclude only those vaccinated within two weeks of initial dosing, or those with plans to be vaccinated within two weeks after completion of dosing.

### Study Endpoints

The primary endpoint for the trial was the safety and tolerability of IN-006. This was assessed by monitoring treatment-emergent adverse events, pre- and post-dose vital signs, ECG, FEV_1_, SpO_2_, hematology and chemistry safety blood tests, and physical examinations. Follow-up continued for 28 days, with assessments on the days indicated in Figure 1B. Exploratory outcomes were drug levels in nasal fluid and serum pre-dose and at intervals post-dose.

### Randomization and Blinding

A randomization schedule was prepared using validated software (SAS) by statistical team members who had no responsibility for monitoring and data management of this study, with provisions for each sentinel pair to include one active and one saline placebo assignment, and for the overall ratio of active to placebo assignment of each cohort to be 3:1. The randomization code was held by unblinded pharmacy staff who prepared the doses in matching syringes with identical appearances for loading into the nebulizer by clinical staff.

### Interventions/Clinical Procedures

IN-006 drug substance was produced under Good Manufacturing Practices (GMP) and supplied as a liquid formulation in glass vials from the manufacturer, Celltrion, Inc. IN-006 was provided in a syringe to be loaded into the InnoSpire Go vibrating mesh nebulizer (Koninklijke Philips N.V., cleared under 510K K170853). Placebo participants received an identical syringe containing saline instead of IN-006.

Participants were instructed to breathe in slowly through the nebulizer mouthpiece and to breathe out through their nose. Nasal fluid was obtained by rotating a flocked swab (Copans Cat. # 56380CS01) for 10-15 seconds at mid-turbinate depth (4-5 cm). Sampling alternated between right and left nostrils during sequential sample collection timepoints. The amount of nasal fluid sample collected by each individual swab was determined by weighing the sample-containing swab and sample tube before and after it was incubated in buffer for extraction, rinsed, and oven dried. Nasal concentrations are therefore reported as ng IN-006 per gram of nasal fluid, which can be approximately interpreted as ng/mL. Sampling times for nasal fluid and serum are shown in Figure 1B. Vital signs and FEV_1_ were measured before nebulization and 15 and 30 minutes after completion of nebulization.

### Measurement of IN-006 concentration in human serum and nasal fluid, and PK modeling

Please refer to the online supplement for further details on sample extraction, ELISA design, and PK modeling.

### Statistical analysis

Sample size was chosen according to conventions for Phase 1, first-in-human studies. Formal sample size and power calculations were not performed. Continuous variables were summarized using descriptive statistics including number of non-missing observations, mean, SD, median, minimum, and maximum values. Categorical variables were summarized with frequency counts and percentages. Placebo recipients in different cohorts were pooled. The safety analysis included all randomized participants who received any dose of study drug. The pharmacokinetic population included all participants who received any dose of IN-006. No inferential statistical tests were conducted. Serum PK parameters of IN-006 were determined using Phoenix WinNonlin version 8.3.

## Results

From among 102 adults who were screened, 23 participants were sequentially assigned to one of three cohorts. The first participant was randomized on September 22, 2021, and the last participant visit was on December 29, 2021. Of these participants, 17 were randomly assigned to receive IN-006, and 6 were randomly assigned to receive placebo. All 23 participants received their assigned treatment as intended and completed the final study visit on Study Day 29. The study was completed on December 29, 2021. Participant flow is diagrammed in Figure 1A, and participant demographics are listed in Table 1.

**Table 1:** Demographics of enrolled participants.

|  | **IN-006 SD**<br>**Low**<br>**(n=5)** | **IN-006 SD**<br>**High**<br>**(n=6)** | **IN-006 MD**<br>**(n=6)** | **Placebo**<br>**(n=6)** | **Overall**<br>**Total**<br>**(n=23)** |
| --- | --- | --- | --- | --- | --- |
| **Sex** |  |  |  |  |  |
| Female | 1 (20%) | 3 (50%) | 2 (33.3%) | 3 (50%) | 9 (39.1%) |
| Male | 4 (80%) | 3 (50%) | 4 (66.7%) | 3 (50%) | 14 (60.9%) |
| **Age, years** |  |  |  |  |  |
| Mean | 30.0 | 33.0 | 24.2 | 35.7 | 30.7 |
| Range | 21-37 | 20-50 | 18-31 | 28-52 | 18-52 |
| **Race** |  |  |  |  |  |
| White | 3 (60%) | 4 (66.7%) | 4 (66.7%) | 5 (83.3%) | 16 (69.6%) |
| Black or African American | 0 | 0 | 0 | 0 | 0 |
| Asian | 2 (40%) | 2 (33.3%) | 1 (16.7%) | 1 (16.7%) | 6 (26.1%) |
| American Indian or Alaska Native | 0 | 0 | 1 (16.7%) | 0 | 1 (4.3%) |
| Native Hawaiian or other Pacific Islander | 0 | 0 | 0 | 0 | 0 |
| Australian Aboriginal or Torres Strait Islander | 0 | 0 | 0 | 0 | 0 |
| Other | 0 | 0 | 0 | 0 | 0 |
| **Ethnic Origin** |  |  |  |  |  |
| Not Hispanic or Latino | 5 (100%) | 6 (100%) | 5 (83.3%) | 5 (83.3%) | 21 (91.3%) |
| Hispanic or Latino | 0 | 0 | 1 (16.7%) | 1 (16.7%) | 2 (8.7%) |
| **Weight (kg)** |  |  |  |  |  |
| Mean | 83.7 | 68.7 | 68.5 | 85.2 | 76.2 |
| Range | 53.9-111 | 53-99.8 | 50.8-87.6 | 70.3-101.8 | 50.8-111 |

### Safety and Tolerability

Treatment emergent adverse events (TEAEs) are listed in Table 2. Nebulization of IN-006 was well-tolerated and completed in an average of 6 minutes for the 90 mg dose (range 4-9 minutes). Eight (53.3%) of the 15 participants included in the single ascending dose cohorts experienced at least 1 TEAE (6 of 11 receiving IN-006, 2 of 4 receiving placebo). Among the 11 participants receiving IN-006, the most frequently reported TEAEs in the SAD cohorts were headache (2/11; 18.2%) and oropharyngeal pain (2/11; 18.2%). All but 1 TEAE were mild. One participant who received IN-006 low dose (30 mg) experienced a moderate event (increased transaminases on Day 29), which was not considered to be related to study drug by the investigator. Three (3/15; 20.0%) participants experienced at least 1 TEAE considered related to study drug by the investigator. These events included headache, cough, and oropharyngeal pain. All 3 related TEAEs were mild and resolved. There was no evidence of a dose-related effect.

**Table 2:**
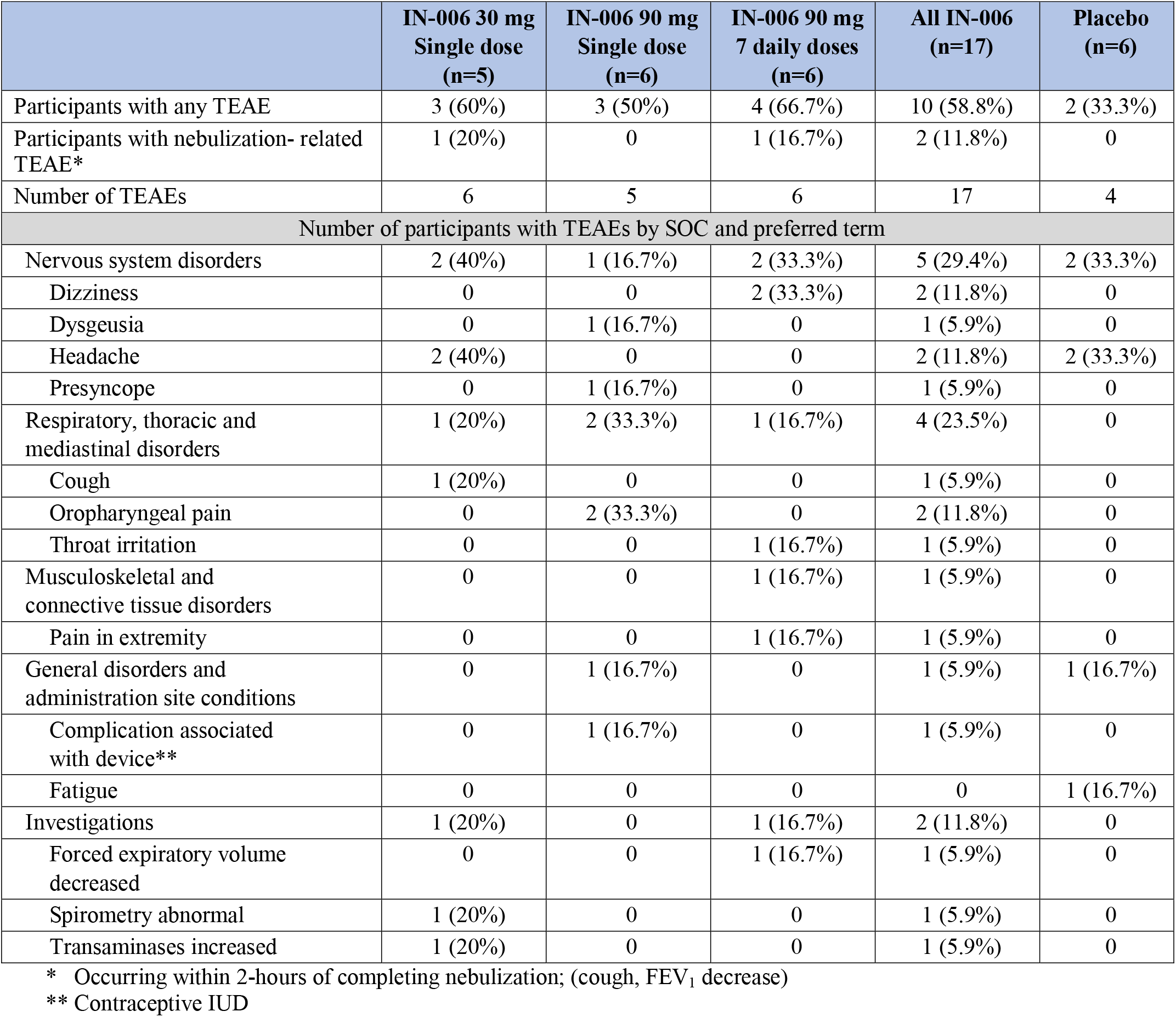
Summary of Adverse Events.

**Table 3:** Summary of Key Intranasal PK Parameters by Cohort and Dose Day.

| **Cohort /**<br>**Dose Day**<br>**Treatment** | **Mean Intranasal Swab**<br>**(CV%)** |  |
| --- | --- | --- |
|  | **C<sub>3h after dose</sub>**<br>**[µg/g]** | **AUC**<br>**[hr\*µg/g]** |
| SAD Cohort 1<br>30 mg IN-006 single dose, n=5 | 260.58<br>(130%) | 3,149.01<br>(129%) |
| SAD Cohort 2<br>90 mg IN-006 single dose, n=6 | 710.17<br>(128%) | 10,377.44<br>(123%) |
| MD Cohort 3, Day 1<br>90 mg IN-006 once daily, n=6 | 921.17<br>(76%) | 10,230.55<br>(76%) |

C<sub>3h after dose</sub> : Concentration 3 hours after dose

**Table 4:** Summary of Key Serum PK Parameters by Cohort and Dose Day.

| Cohort | Median<br>(range) | Mean Serum<br>(%CV) |  |  |
| --- | --- | --- | --- | --- |
| | $t_{\max}$<br>(hr) | $C_{\max}$<br>(ng/mL) | AUC<sub>0-tlast</sub><br>(hr*ng/mL) | $t_{1/2}$<br>(hr) |
| SAD Cohort 1<br>30 mg IN-006, n=5 | 121.5<br>(72 – 122) | 71.5<br>(62%) | 19,118<br>(97%) | 253<sup>‡</sup><br>(15%) |
| SAD Cohort 2<br>90 mg IN-006, n=6 | 118.9<br>(116 – 121) | 88.9<br>(56%) | 27,624<br>(82%) | 292<sup>‡</sup><br>(27%) |
| MD Cohort 3<br>90 mg IN-006 once daily, n=6 | 188.4<br>(145-216) | 580.3<br>(41%) | 257,170*<br>(37%) | 401.7<br>(36%) |

<sup>‡</sup> n = 2 for samples with a sufficient number of samples with detectable concentrations
\* For Cohort 3, the value of 257,170 represents the AUC calculated through tlast, to allow for direct comparison to Cohorts 1 and 2. However, since Cohort 3 had more quantifiable samples at later timepoints, we were also able to calculate AUC until the serum concentration theoretically hit zero (AUC<sub>infinity\_obs</sub>), which was 404,034 hr\*ng/mL, with %CV of 35%. The AUC<sub>infinity</sub> was not calculated for Cohorts 1 and 2 because too many samples were BLQ at the later timepoints, making the estimate for the terminal phase of elimination less reliable (see online supplement).

In the multiple dose cohort, no TEAEs were reported in participants receiving placebo. Among the 6 participants receiving IN-006, 4 (66.7%) participants experienced at least 1 TEAE. The most frequently reported TEAE was dizziness (2/6; 33.3%). All but 1 TEAE were mild. One participant receiving IN-006 experienced a moderate event (pain in extremity), which was considered unlikely to be related to study drug by the investigator. Two (33.3%) participants experienced at least 1 TEAE considered related to study drug by the investigator. These drug-related TEAEs were dizziness and decrease in FEV_1_: the latter was noted 15 minutes after nebulization, was not associated with symptoms or abnormal vital signs, resolved within 15 minutes, and did not recur with subsequent doses. Both events were mild.

No severe TEAEs, SAEs, or TEAEs leading to discontinuations were reported in either the single dose or multiple dose cohorts. The most frequently reported TEAEs in participants receiving IN-006 across all three cohorts were headache (2/23; 11.8%) and oropharyngeal pain (2/23; 11.8%), both appearing only in the SAD cohorts and not in the multiple dose cohort. There were no unexpected safety signals.

### Pharmacokinetics

For single dose cohorts, the mean concentrations of IN-006 per gram of nasal fluid were 261 µg/g and 710 µg/g for the 30 mg and 90 mg dose, respectively, measured 3 hrs after dosing; the difference in these values is consistent with a 3-fold increase in the dose administered. In the multiple dose cohort, the repeated dosing provided additional opportunities for more nasal concentration measurements across more time points. The nasal concentrations measured 30 mins after dosing on Days 1, 2, and 3 averaged 773 µg/g, which was higher than the average concentration measured 3 hrs after dosing (388 µg/g). This indicates that peak exposure occurred very shortly after dosing, and the nasal concentrations had appreciably reduced by 3 hours post-dose. There was minimal intranasal accumulation upon repeated dosing, as the concentrations of IN-006 measured 22 hrs after a single dose were <2% of the concentrations immediately following dosing (Figure 2). The difference in nasal concentrations between 30 mins and 22 hrs post-dose suggest the interval in the single dose groups represented ∼4-9 half lives, and the difference between concentrations at 30 mins and 3 hrs post-dose was roughly half. Both findings are consistent with an intranasal half-life on the order of 2-6 hours, markedly longer than the timescale of mucociliary clearance transit time estimates of ∼5-15 minutes from saccharin transit time tests [16, 17].

**Figure 2:**
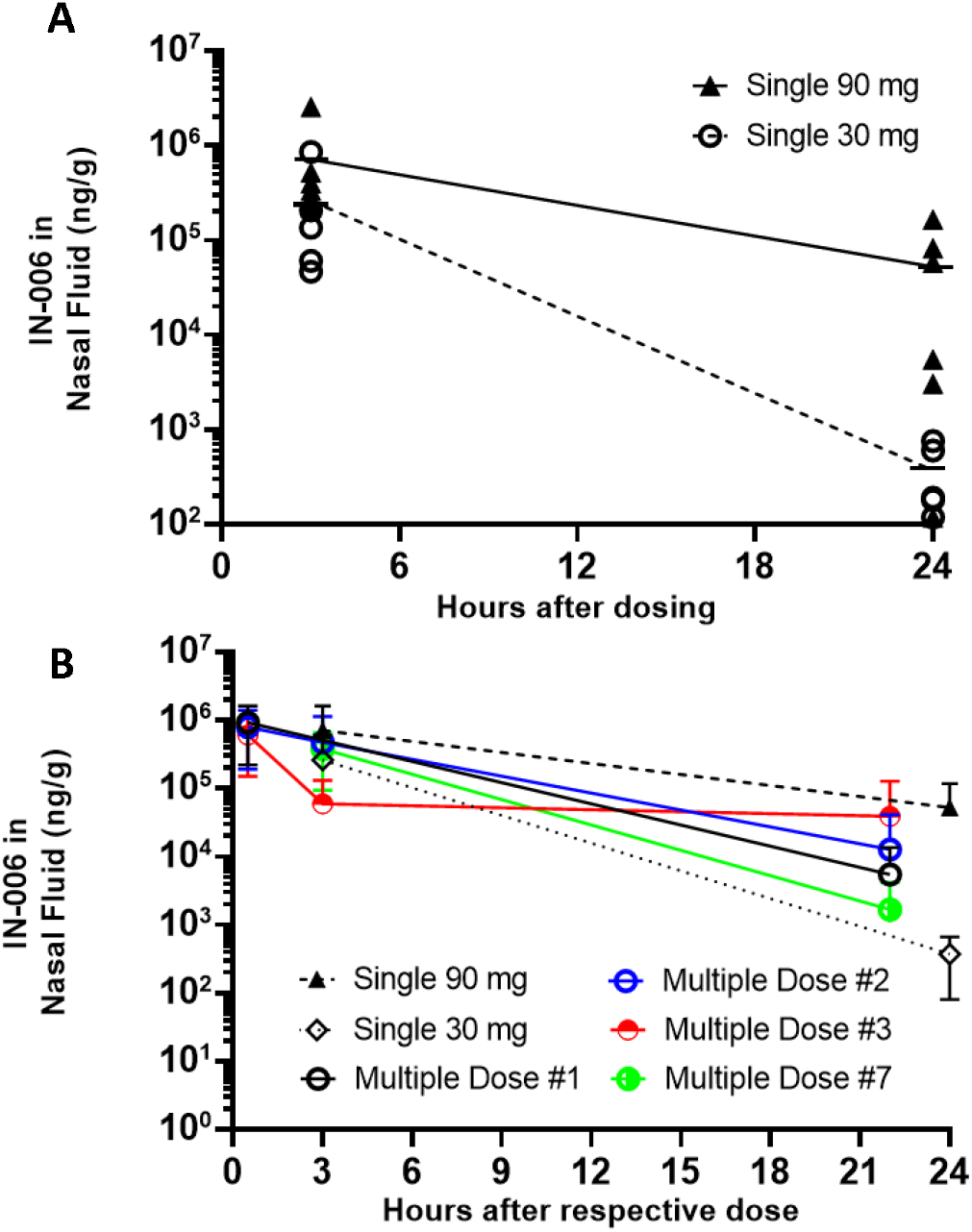
**(A)** Nasal fluid concentrations in single dose cohorts. **(B)** Comparison of nasal concentrations between single dose and multiple dose cohorts at varying times after each dose. LLOQ varied by sample, depending on the mass of nasal fluid collected on swab. For this figure, for samples that were measured as BLQ, concentration was reported as the midway point between 0 and LLOQ for the purpose of calculating group average concentrations at each timepoint. For example, a BLQ sample with an LLOQ of 1,000 ng/mL was calculated at 500 ng/mL. For panel B, please refer to online supplemental figure 1 for further details and for an updated version of this figure in which two datapoints suspected to be swapped at the time of data collection (one individual in Multiple Dose 3, at 3h and 22h) were corrected. Data here in Panel B is are not corrected for this the suspected error.

**Figure 3:**
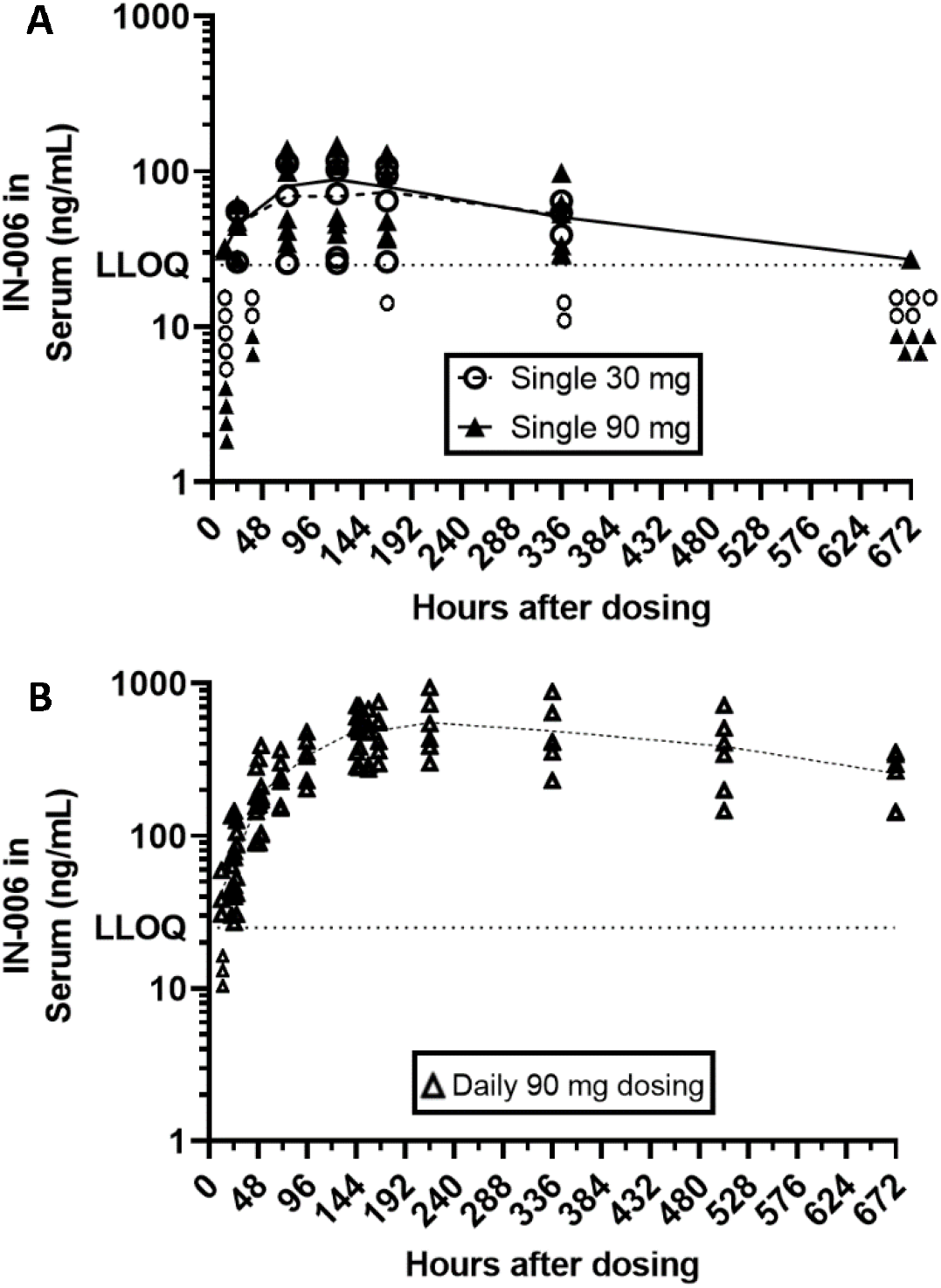
Serum IN-006 concentrations in **(A)** single dose cohorts, and **(B)** multiple dose cohort (last dose administered at 144 h). Symbols plotted below the dashed LLOQ line at 25 ng/mL represent the number of samples in each group that were BLQ at each timepoint.

Serum concentrations of IN-006 were detectable by 12 hrs following nebulization at the 90 mg dose and continued to rise through 120 hrs after a single dose (cohorts 1 and 2), or through 216 hrs following the first dose in those who received multiple doses (cohort 3). In the multiple dose cohort, Cmax in the serum occurred following the final dose, indicating accumulation. The elimination half-life of IN-006 in the serum was estimated to be ∼253, 292, and 402 h in the 30 mg single dose, 90 mg single dose, and the 7 daily 90 mg dose cohorts respectively, comparable with the previously estimated elimination half-life of regdanvimab from the serum following intravenous administration (288 h [18]). Although the serum concentrations of IN-006 were markedly lower than those in the nasal fluid (Serum C_max_ of 0.52 µg/mL, compared to 990 µg/mL in nasal fluid), they were still significantly greater than the IC_50_ of IN-006 (∼0.01 µg/mL) against susceptible variants.

## Discussion

A longstanding dogma has been that it is highly challenging to stably nebulize mAbs [19-22], and that biologic drugs would be quickly eliminated from the respiratory tract either by systemic absorption, physical mucociliary clearance, or degradation by alveolar macrophages, making it difficult to sustain therapeutic concentrations [23, 24]. However, in this Phase 1 study, we found that IN-006, a reformulation of regdanvimab for nebulized delivery at point of care, was safe and well tolerated in healthy adults. High concentrations of drug were recovered from nasal samples, as well as appreciable levels detected in the serum. We also found that the treatment was easily self-administered by participants and was completed within minutes, with minimal side effects. Encouragingly, the mean concentrations of IN-006 measured in nasal secretions, representing IN-006 deposited in the nasal passages after being exhaled from the lungs, were well above its IC_50_, even 22-24 hrs after dosing. In the multiple dose cohort that received seven daily 90 mg doses, we observed a mean nasal fluid IN-006 concentration of 920 µg/mL 30 minutes after the initial dose and a mean concentration of 5.8 µg/mL 22 hours later, prior to receiving a second dose. The ability to maintain mAb concentrations that ranged from 3-7 orders of magnitude higher than the IC_50_ for regdanvimab and other COVID mAbs against susceptible variants (∼4-20 ng/mL, [13, 25]) strongly support our proposed once-daily dosing regimen. Since SARS-CoV-2 infection and replication initiates in the upper respiratory tract, our efficient delivery of IN-006 to the nasal passages suggests it may provide a highly effective treatment for mild to moderate COVID-19 that allows earlier resolution of the infection and reduced risk of progression to severe COVID.

Although mAbs have proven to be effective therapeutics for COVID-19, the necessity for administration by IV, IM, or SC routes has limited the scope of their use in clinical practice. The requirement for infusion centers and post-dosing observation for intravenous administration have severely limited the number of patients that have received treatment and greatly increased costs [8, 9]. IM injections, although shortening administration time, are limited by the volume that can be administered per injection (∼5 mL) [26], which in turn limits the dose of mAb that can be dosed per injection and can be painful when maximum injection volumes are used. In contrast, nebulized delivery using a handheld nebulizer enables the convenience of at-home dosing and takes only minutes to complete. Furthermore, IV, IM, and SC routes provide mAb Cmax to the airway lining fluid from the blood only after a delay of one or more days, and, even then, only achieve airway concentrations that are a fraction of the concentrations in plasma [10, 11, 14]. For instance, in a recent clinical trial of the anti-influenza mAb CR6261 given as a single 50 mg/kg dose IV, the peak nasal concentration was not achieved until 2 days after IV infusion [12] and the peak nasal concentration of 0.597 µg/mL was ∼10-fold lower than the concentrations we observed for IN-006 at the trough of our daily inhaled dosing (∼5.8 µg/mL), despite the much lower total dose of IN-006 compared to CR6261 (90 mg IN-006 vs. ∼2,000-4,000 mg CR6261 [12]). We believe that the increased convenience and more efficient pulmonary delivery will likely make inhalation the preferred route of mAb delivery for treating acute respiratory infections.

Serious disease due to SARS-CoV-2 is accompanied by the spread of the virus from the site of the initial upper respiratory tract infection to the deep lungs [7]. Unfortunately, the exact timing of such spread is likely to be highly variable between individuals. Indeed, there is evidence suggesting viruses can reach the LRT even during the early stages of disease, around the time that symptoms emerge [27, 28]. Thus, we believe dosing to both the URT and LRT, rather than focusing exclusively on the URT (e.g., via nasal sprays), will be important to broaden the treatment window and reduce risk of COVID-induced pneumonia and hospitalization. While IN-006 levels in the LRT were not directly measured in this study, the appreciable serum concentrations and the delayed serum Tmax both strongly suggest we are efficiently delivering IN-006 into the LRT and the deep lung. Indeed, in a toxicokinetic multiple dose nebulization study of IN-006 in rats, IN-006 concentrations in airway fluid exceeded the serum concentrations by ∼100-fold [14]. Efficient delivery into the LRT is a direct consequence of our design requirement for the vibrating mesh nebulizer. The droplet sizes generated by the nebulizer (the fine particle fraction, i.e. droplets <5 um, and particularly those <2.5 um [15]) were intentionally selected to deliver a portion of the mAbs throughout the LRT and deep lung. Furthermore, the fact that we observed a slow steady rise of serum concentrations in single dose cohorts over ∼4 days, and peak serum concentrations in the multiple dose cohort over ∼9 days (or 2 days after last dose), implies that we are sustaining high levels of IN-006 in the deep lungs for at least 2-4 days after a single dose (or after the final dose). Assuming a ratio of 100:1 of antibody concentrations in the airway fluid:serum, the mean serum concentration of 550 ng/mL at Day 9 should translate to pulmonary concentrations on the order of 55 µg/mL, which is >3 orders of magnitude above the IC_50_, and comparable to the serum concentrations achieved with some IV/IM-dosed mAbs. The very high mAb levels sustained relative to the intrinsic activity of the mAb (IC_50_) may continue to provide effective treatment against variants, even in the presence of appreciable genetic drift, and may reduce the risk of viral escape. This also suggests that shorter durations of therapy may afford appreciable protection against hospitalization.

Despite significant extrapulmonary manifestations of severe COVID-19, and despite frequent detection of SARS-CoV-2 RNA in blood, infectious SARS-CoV-2 is rarely detected in the blood of infected patients [29, 30], suggesting that extrapulmonary disorders are in many cases caused by indirect factors such as the inflammatory response rather than extrapulmonary viral infection. Nonetheless, we find it reassuring to observe that mean serum levels of IN-006 achieved after nebulized delivery were in excess of its IC_50_ by at least one order of magnitude.

Regdanvimab (administered IV) was shown to be highly efficacious for preventing severe COVID-19 in a global Phase 3 study, leading to its approval in Republic of Korea and European Union (EMEA/H/C/005854) for preventing severe disease in patients presenting with mild to moderate COVID-19, and emergency use authorization (EUA) or conditional marketing authorization in several additional countries worldwide. Nevertheless, as SARS-CoV-2 continues to evolve, we are pursuing combining IN-006 with a second potent neutralizing mAb to create a mAb cocktail that possesses potent binding activity against every variant tested to date, including Omicron BA.4/5. In light of the surprisingly long airway retention of IN-006 observed here, we plan to reduce the duration of dosing from 7 days to 5 days in future clinical studies, offering even greater convenience to the patients.

Our study had several limitations. The population we studied was small, as is typical of first-in-human studies. Moreover, only healthy adults were enrolled in our study, limiting its generalizability. Determining the full safety profile of nebulized IN-006 will require additional studies in larger numbers of more diverse individuals. The low recovered sample mass from nasal swabs required substantial dilution for sample recovery, limiting the sensitivity of the assay. As a result, some 24-hour post-dose samples and all later nasal samples were below the limit of quantification. Direct determination of drug levels achieved in the deep lung was beyond the scope of this study, and additional studies with bronchioalveolar lavage are contemplated in the further development of IN-006.

In conclusion, IN-006, a reformulation of regdanvimab for inhaled delivery, was found to be safe and well tolerated in healthy participants at single doses of 30 mg and 90 mg, as well as seven consecutive daily doses of 90 mg. Nebulization resulted in IN-006 levels in nasal fluids, and likely the lungs, that were orders of magnitude above the inhibitory concentrations of sensitive SARS-CoV-2 variants within 30 minutes, and the continued rise of serum concentration for days after dosing implied substantial lasting IN-006 levels in the lungs. These data support the ongoing clinical development of IN-006 as an inhaled treatment for COVID-19.

## Data Availability

Data produced in the present study are available upon reasonable request to the authors

## References

1. Pinto, A.L., et al., Ultrastructural insight into SARS-CoV-2 entry and budding in human airway epithelium. Nature Communications, 2022. 13(1): p. 1609.

2. Lee, I.T., et al., ACE2 localizes to the respiratory cilia and is not increased by ACE inhibitors or ARBs. Nature Communications, 2020. 11(1): p. 5453.

3. Jia, H.P., et al., ACE2 receptor expression and severe acute respiratory syndrome coronavirus infection depend on differentiation of human airway epithelia. J Virol, 2005. 79(23): p. 14614–21.

4. Zhang, L., et al., Respiratory syncytial virus infection of human airway epithelial cells is polarized, specific to ciliated cells, and without obvious cytopathology. J Virol, 2002. 76(11): p. 5654–66.

5. Sims, A.C., et al., SARS-CoV replication and pathogenesis in an in vitro model of the human conducting airway epithelium. Virus Res, 2008. 133(1): p. 33–44.

6. Zhang, L., et al., Infection of ciliated cells by human parainfluenza virus type 3 in an in vitro model of human airway epithelium. J Virol, 2005. 79(2): p. 1113–24.

7. Hou, Y.J., et al., SARS-CoV-2 Reverse Genetics Reveals a Variable Infection Gradient in the Respiratory Tract. Cell, 2020. 182(2): p. 429-446.e14.

8. Lai, S.K., M.D. McSweeney, and R.J. Pickles, Learning from past failures: Challenges with monoclonal antibody therapies for COVID-19. J Control Release, 2021. 329: p. 87–95.

9. Cruz-Teran, C., et al., Challenges and opportunities for antiviral monoclonal antibodies as COVID-19 therapy. Adv Drug Deliv Rev, 2021. 169: p. 100–117.

10. Hart, T.K., et al., Preclinical efficacy and safety of mepolizumab (SB-240563), a humanized monoclonal antibody to IL-5, in cynomolgus monkeys. J Allergy Clin Immunol, 2001. 108(2): p. 250–7.

11. Dall’Acqua, W.F., P.A. Kiener, and H. Wu, Properties of human IgG1s engineered for enhanced binding to the neonatal Fc receptor (FcRn). J Biol Chem, 2006. 281(33): p. 23514–24.

12. Han, A., et al., Safety and Efficacy of CR6261 in an Influenza A H1N1 Healthy Human Challenge Model. Clin Infect Dis, 2020.

13. van der Straten, K., et al., Optimization of Anti-SARS-CoV-2 Neutralizing Antibody Therapies: Roadmap to Improve Clinical Effectiveness and Implementation. Front Med Technol, 2022. 4: p. 867982.

14. McSweeney, M., et al., Stable nebulization and muco-trapping properties of Regdanvimab/IN-006 supports its development as a potent, dose-saving inhaled therapy for COVID-19. bioRxiv, 2022: p. 2022.02.27.482162.

15. Darquenne, C., Aerosol deposition in health and disease. Journal of aerosol medicine and pulmonary drug delivery, 2012. 25(3): p. 140–147.

16. Emma, R., et al., Short and Long Term Repeatability of Saccharin Transit Time in Current, Former, and Never Smokers. Front Physiol, 2020. 11: p. 1109.

17. Rodrigues, F., et al., Particularities and Clinical Applicability of Saccharin Transit Time Test. Int Arch Otorhinolaryngol, 2019. 23(2): p. 229–240.

18. Syed, Y.Y., Regdanvimab: First Approval. Drugs, 2021. 81(18): p. 2133–2137.

19. Respaud, R., et al., Nebulization as a delivery method for mAbs in respiratory diseases. Expert Opin Drug Deliv, 2015. 12(6): p. 1027–39.

20. Mayor, A., et al., Inhaled antibodies: formulations require specific development to overcome instability due to nebulization. Drug Deliv Transl Res, 2021. 11(4): p. 1625–1633.

21. Bodier-Montagutelli, E., et al., Protein stability during nebulization: Mind the collection step! Eur J Pharm Biopharm, 2020. 152: p. 23–34.

22. Bodier-Montagutelli, E., et al., Designing inhaled protein therapeutics for topical lung delivery: what are the next steps? Expert Opin Drug Deliv, 2018. 15(8): p. 729–736.

23. Loira-Pastoriza, C., J. Todoroff, and R. Vanbever, Delivery strategies for sustained drug release in the lungs. Adv Drug Deliv Rev, 2014. 75: p. 81–91.

24. Suri, R., The use of human deoxyribonuclease (rhDNase) in the management of cystic fibrosis. BioDrugs, 2005. 19(3): p. 135–44.

25. Liu, L., et al., Striking antibody evasion manifested by the Omicron variant of SARS-CoV-2. Nature, 2022. 602(7898): p. 676–681.

26. Nagy, C.F., et al., Safety, Pharmacokinetics, and Immunogenicity of Obiltoxaximab After Intramuscular Administration to Healthy Humans. Clinical pharmacology in drug development, 2018. 7(6): p. 652–660.

27. Chen, P.Z., et al., SARS-CoV-2 shedding dynamics across the respiratory tract, sex, and disease severity for adult and pediatric COVID-19. eLife, 2021. 10: p. e70458.

28. Lim, A.-Y., et al., Modeling the early temporal dynamics of viral load in respiratory tract specimens of COVID-19 patients in Incheon, the Republic of Korea. International Journal of Infectious Diseases, 2021. 108: p. 428–434.

29. Wang, W., et al., Detection of SARS-CoV-2 in Different Types of Clinical Specimens. Jama, 2020. 323(18): p. 1843–1844.

30. Ram-Mohan, N., et al., SARS-CoV-2 RNAemia Predicts Clinical Deterioration and Extrapulmonary Complications from COVID-19. Clin Infect Dis, 2022. 74(2): p. 218–226.

31. Celltrion Submits Investigational New Drug (IND) Application to Initiate a Global Phase III Clinical Trial Evaluating an Inhaled COVID-19 Antibody Cocktail Therapy. 2022.

