## Supplemental Figures and Methods for "A randomized, double-blind, Phase 1 study of IN-006, an inhaled antibody treatment for COVID-19"

*Measurement of IN-006 concentration in human serum and nasal fluid*

Non-GLP ELISA-based bioanalytical methods were used for the analysis of IN-006 in human serum and human nasopharyngeal fluid. These assays used anti-idiotypic antibodies that bound IN-006 for capture and detection. Reference mAb and anti-ID antibodies, 18D2 and 18E5, were provided by Celltrion, Inc. The streptavidin-HRP conjugate of the anti-ID 18E5 was made using a Lightning Link® HRP Conjugation Kit (Abcam, Cat# ab103890). For the nasopharyngeal samples, the initial step involved extraction of IN-006 from nasopharyngeal swab prior to dilution to run on the ELISA. No extraction step was necessary for the serum samples. IN-006 standards, human serum samples, and quality control samples were diluted and added to a microtiter assay plate that had been coated with anti-ID 18D2, and the plate was then blocked. After washing, the streptavidin-HRP conjugated anti-ID 18E5 was then added, incubated, and washed. The plate was developed using 3,3',5,5',-tetramethyl benzidine (TMB, Thermoscientific, Cat# 30429) and stopped with 2N sulfuric acid (Fisher, Cat# SA212-4). The absorbance was measured at 450 nm and 650. Data analysis was performed by creating a dose-response curve for the reference standards on each plate by plotting the signal in each well (y -axis) against the corresponding concentration of IN-006 (x-axis). The response plot was then fit with a 4-parameter logistic (4PL) non-linear regression model (using SoftMax software). The lower limit of quantitation (LLOQ) for the serum assays was 5 ng/mL, and the samples were diluted at least 5-fold, resulting in an effective LLOQ of 25 ng/mL. The LLOQ for the nasopharyngeal samples was variable (between 105-1,770 ng/g) due to the variability in the mass of collected samples and the extent of subsequent dilution of the nasopharyngeal fluid.

*Nasal PK analysis.*

Nasal fluid pharmacokinetic data were available for all participants for all planned sampling time points. Mean IN-006 concentration in nasal fluid over time is plotted for each cohort. In each Cohort, C_max_ in the nasal fluid was observed at the first sampling point collected following dosing. In the single-dose Cohorts and multiple dose Cohort, a true estimate of elimination rate was not able to be determined due to limited availability of samples with measurable drug concentrations (i.e., fewer than 3 concentration estimates after t_max_), but rough estimates of half-life are described in the main text. Intranasal PK parameters of IN-006 were determined using Phoenix WinNonlin version 8.3. All times used in the calculation of pharmacokinetic parameters were the actual elapsed time from the most recent treatment administration, with the exception of pre-dose data which was given the nominal time of 0.00 h.

Each nasal swab sample had a different amount of nasal fluid that was able to be extracted, and therefore each sample had a different dilution factor prior to ELISA assay, leading to sample-specific LLOQs. For the calculation of the average nasal swab concentrations shown in Figure 2, samples that returned results that were BLQ upon ELISA were assumed to have a concentration halfway between zero and the sample-specific LLOQ. Compared to either inputting zero or the LLOQ value for each BLQ point, we found that this midway assumption did not have a substantial impact on group averages or result interpretations.

*Serum PK analysis.*

Serum pharmacokinetic data were available for all participants for all planned sampling time points. Mean IN-006 concentration in serum vs. time is plotted for each cohort in Figure 3. Key parameters of systemic exposure (t_max_, C_max_, AUC_0-tlast_, and t_1/2_, also AUC_0-infinity_obs_ for MD) is presented in Table 4, showing median and range for t_max_ and mean (%CV) for C_max_ and AUC. In the single-dose Cohorts, a valid estimate of elimination rate was able to be determined for only 2 of 5 participants in Cohort 1 and for only 2 of 6 participants in Cohort 2 due to limited availability of samples with measurable drug concentrations (i.e., fewer than 3 concentration estimates after t_max_). In the MD Cohort, however, estimates of elimination half-life could be generated for all participants, with a mean span of 0.97 and an R^2^ of ~0.95, suggesting that the collected samples described the terminal log-linear phase of elimination well, allowing the calculation of AUC_0-infinity_obs_. For Cohorts 1 and 2, the last timepoint collected had many samples that were BLQ, decreasing the span for calculation of elimination rate constant and therefore decreasing certainty in AUC_0-infinity_ predictions, so AUCs were simply reported through tlast in Table 4.

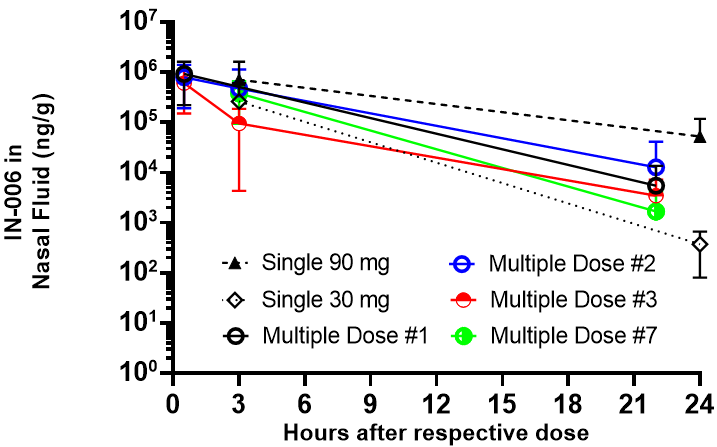

**Supplemental Figure 1**: Replacement panel for main text Figure 2B with updated datapoints for red group. For the data group of Multiple Dose 3, shown in red, the concentrations measured at 3 hours and 22 hours for one subject have been switched, to correct for a suspected change of samples during collection. As recorded in the main text, one individual in the Multiple Dose group after Dose 3 had an apparently higher concentration at 22h following the dose than 3h following the dose, which was the exact reverse of the trends for every other individual in every group tested. Suspecting that these two datapoints (or samples) had unintentionally been switched, we have switched those two datapoints for the group shown in red to reflect what we believe to have truly occurred. This would reflect the expected decline from 3h to 22h after Multiple Dose 3.
